# Exposure patterns and the risk factors of zoonotic Crimean Congo hemorrhagic fever virus amongst pastoralists, livestock and selected wild animals species at the human/livestock/wildlife interface in Isiolo County, upper eastern Kenya

**DOI:** 10.1101/2024.03.20.24304573

**Authors:** Eugine Mukhaye, James Akoko, Richard Nyamota, Athman Mwatondo, Mathew Muturi, Daniel Nthiwa, Lynn J. Kirwa, Joel L. Bargul, Hussein M. Abkallo, Bernard Bett

## Abstract

Crimean Congo hemorrhagic fever (CCHF) is a tick-borne zoonotic disease caused by CCHF virus (CCHFV), and has a complex transmission cycle that involves a wide range of hosts including mammalian and some species of birds. We implemented a seroepidemiological study in Isiolo County, Kenya, to determine relative seroprevalences of CCHFV in pastoralists, livestock and in wild animals’ species. In addition, we identified subject and environment level factors that influence exposure to CCHFV. Pastoralists (n = 580) and livestock (n = 2,137) were recruited into the study through a multistage random sampling technique, and addition, various species of wild animals (n = 87) were also sampled conveniently. Serum samples from all recruited subjects were collected and screened for CCHFV antibodies using ID.vet multispecies, double-antigen IgG enzyme-linked immunosorbent assay (ELISA). The overall anti-CCHFV IgG seroprevalences in pastoralists, cattle, goats, sheep and camels were 7.2% [95% CI: 3.1–15.8%], 53.9% [95% CI: 30.7–50.9%], 11.6% [95% CI: 7.2–22.5%], 8.6% [95% CI: 3–14%] and 89.7% [95% CI: 78–94%], respectively. On average, wild animals’ hosts had CCHFV seroprevalence of 41.0% [95% CI: 29.1–49.4%], and giraffes had the highest seroprevalence of 75.0% [95% CI: 63.2–79.3%]. Analysis using mixed effects logistic regression models showed that CCHFV exposure in pastoralists was significantly associated with male gender, being over 30 years and belonging to a household with a seropositive herd. In livestock, mature animals, high normalized difference vegetation indices and high vapour pressure deficit were significantly associated with CCHFV exposure. Age, sex and species of wild animals were considered as the risk factors in the analysis, but none of these variables was significant (P-value = 0.891, 0.401 and 0.664 respectively). A look at the risk factors suggests that the environmental factors such as the NDVI (P-value = 0.032) and the vapour pressure deficit (P-value = 0.004) significantly influences CCHFV exposure in livestock, while the presence of infected animals in a household significantly influences pastoralists exposure (P-value = 0.041). CCHF control in livestock, for example by applying acaricides, could minimize the risk of CCHFV exposure to pastoralists. The key findings of this study will guide policymakers in disease control, and emphasize the need for regular surveillance of zoonotic diseases for emergency preparedness, and also contributes to knowledge on One-Health approach for improved health

**Author summary:** Our study focused on understanding the prevalence and risk factors associated with Crimean Congo hemorrhagic fever virus (CCHFV) among pastoralists, livestock, and wild animals in Isiolo County, Kenya. Through a comprehensive seroepidemiological investigation, we found varying seroprevalence rates across different species, with camels exhibiting the highest prevalence. Wild animals, notably giraffes, also displayed significant seroprevalence rates. Factors such as gender, age, and herd seropositivity were identified as significant contributors to CCHFV exposure among pastoralists, while environmental factors like vegetation indices and vapor pressure influenced livestock exposure.

Our findings underscore the intricate interplay between human, animal, and environmental factors in CCHFV transmission dynamics. By elucidating these factors, we provide crucial insights for policymakers to develop targeted interventions and surveillance strategies, emphasizing the importance of a One-Health approach. Implementing control measures in livestock, such as acaricide application, could effectively mitigate CCHFV transmission to pastoralists. Overall, our study contributes to advancing knowledge in zoonotic disease control and underscores the necessity for proactive measures to enhance public health preparedness.

## 1. Introduction

Crimean Congo hemorrhagic fever (CCHF) is a zoonotic disease that is caused by tick-borne CCHF virus (CCHFV) (Atim et al., 2023). The CCHFV is a fragmented, single-stranded RNA virus of the *Bunyaviridae* family, genus *Nairovirus* (Drosten et al., 2003). CCHF is endemic in parts of Africa, the Middle East, Eastern Europe and Asia (Belobo et al., 2021). Domestic animals, wild animals and some avian species serve as reservoirs of CCHFV, hence maintaining disease transmission (Chinikar et al., 2010).

Humans are infected through bites of infected tick or via contact to the CCHF-infected animal tissues. In addition, the possibility of human-to-human infections has been reported (Chinikar et al., 2012). People exposed to CCHFV acquire the disease by undergoing at least four different exposure phases, namely, incubation, pre-hemorrhagic, hemorrhagic, and recuperation phases (Ergonul et al., 2006; Swanepoel & Paweska, 2011; Whitehouse, 2004). In most people, the disease manifests as a transient febrile illness with symptoms such as nausea, myalgia, photophobia, chills, fever, and a severe headache (Kaya et al., 2011). A small fraction of individuals experience a more severe haemorrhagic syndrome when exposed to the virus after 5 to 14 days. This could be due to differences in the mode of exposure, as a tick bites are expected to provide a bigger infectious dose of the virus compared to that from contact with tissues from an infected animal (Kaya et al., 2011).

CCHF is a significant public health threat owing to its high mortality rates of up to 50% in infected humans (Chinikar et al., 2012). The risk of CCHF is further exacerbated by the lack of an efficacious vaccine at present due to lack of a worldwide representative of CCHFV genotypes, as well as virus evolution in the viral targets, which renders the vacines ineffective (Tipih & Burt, 2020), and the inadequate medication choices for the patients, with ribavirin, a broad anti-RNA virus inhibitor being the only approved treatment (Dowall et al., 2017).

CCHFV is spread to animals through tick bites during their blood meal. Livestock infections with CCHF poses huge threat to the livestock sector due to restrictions on trade, transportation, sale or slaughter of livestock during outbreaks, which results in agricultural and economic losses (Sang et al., 2006). It also poses health risks to the value chain actors in livestock marketing chains (meat processing workers, traders, retailers, consumers) since they may be exposed to the CCHFV via handling of infected livestock tissues and/or acquire the disease through infected tick bites as host-attached ticks would be transported to the livestock market.

CCHFV is endemic in many pastoral rangelands where people, livestock and wild animals share common geographical locations (Blanco-Penedo et al., 2021). This interface brings together wild animals, livestock species, diverse species of ticks, and also humans, which provides the necessary conditions for active transmission of CCHFV from the infected animals, which also act as virus reservoirs, to the uninfected ones (Hewson, 2007).

Little is known about the levels of CCHFV exposure in humans, livestock and wild animals that share common environments. In a study that was conducted in the Maasai Mara ecosystem, Nanyuki, and the Ol Pejeta Conservancy in Kenya, a mean CCHFV seroprevalence of 31.5% in cattle, sheep, and goats was estimated (Blanco-Penedo et al., 2021). Another study that was implemented in the same year in Lake Nakuru National Park, Solio Conservancy, Maasai Mara ecosystem, Meru National Park, and Ol Pejeta Conservancy reported CCHFV seroprevalences of 75.3% and 28.1% in buffalo and cattle, respectively (Obanda et al., 2021). Similarly, a mean CCHFV seroprevalence of 5.3% (n = 1958) was reported in cattle, sheep, and goats that were sampled in western Kenya (Nyataya et al., 2020), while another study that sampled animals from slaughterhouses in Nairobi, Kiambu, and Muranga Counties reported a seroprevalence of 4.2% (n = 2330) (Sang et al., 2006). From these studies, the mean CCHFV seroprevalence in livestock ranges from 4.2% to 31.5% in Kenya. In humans, the mean seroprevalences that have been reported ranged from 1.9% to 2.5% (Lwande et al., 2012).

Our study explored CCHFV seroprevalence in pastrolists, livestock species, and wild animals among the largely pastoral community in Garbatulla subcounty, Isiolo, Kenya. Some wild animals samples were collected from Ngare Mara ward, Isiolo, Kenya. The design used in this work fostered the community and public engagement approach as it brought together the professionals from the public and animal health sectors, and the local communities to address the CCHF burden. The study also utilized several environment data including climatic, topographic and geological data while investigating risk factors that influence the spatial distribution of the disease.

## 2. Materials and Methods

### 2.1 Ethics statement

The study received ethical approval from the Institutional Research Ethics Committee of the International Livestock Research Institute (Reference number: ILRI-IREC2020-07) to collect samples from both livestock and humans. Prior to sampling, written consent was obtained from livestock owners and household heads. For individuals below 18 years of age, written permission was acquired from their parents or legal guardians. Adults aged 18 and above provided written consent before being sampled. All aspects of human and animal sampling, as well as data collection, adhered strictly to the standard operating procedures and guidelines outlined in the ethical approval. The wild animals samples, were collected in collaboration with KWS during their routine surveillance and animal translocation activities and as such no ethical approval was required for the work.

### 2.2 The study site

The study was carried out in Isiolo County, upper eastern Kenya (Fig 1). The area falls in an arid to semi-arid ecological zone and is inhabited by the Borana pastoralists. This community keeps cattle, sheep, goats, and camels, which are adjacent with several species of wild animals that usually stray from the two neighbouring national parks, namely the Meru National Park in the southeast and Samburu National Park in the northwest. These parks were used in the study for opportunistic wild animals sampling.

**Fig 1:**
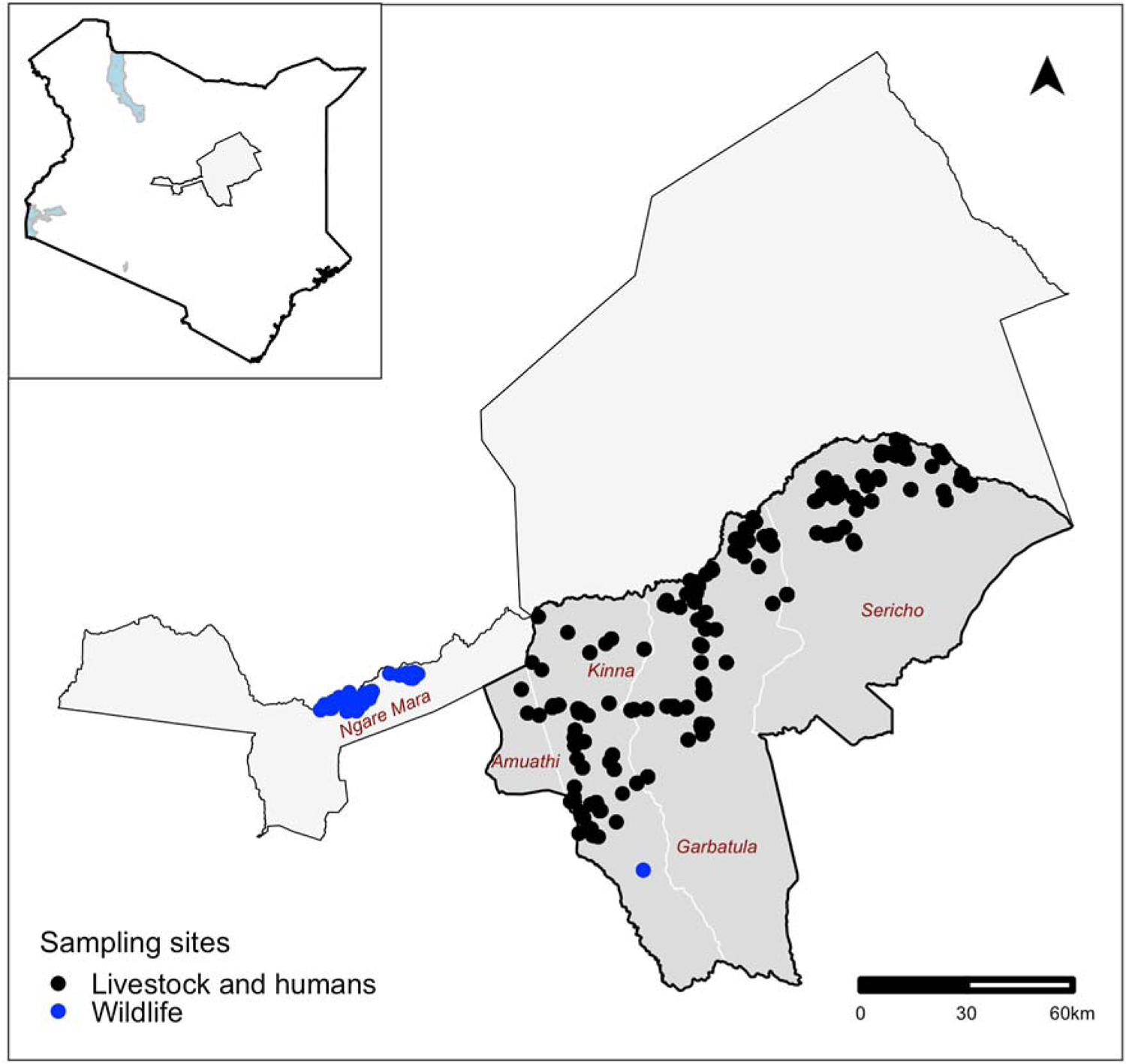
A map showing the sampling site in Isiolo County, Kenya. The black dots represent locations where livestock and human samples were drawn from, while the blue dots represent areas where wild animals were sampled. The background map is available online at the National Geographic World map site (https://www.natgeomaps.com/catalogsearch/result/?q=kenya+map).

The area spans five administrative Wards in the County in a south-west/north-east orientation (Fig 1). This belt was specifically curved out for the study since it represents a unique ecological gradient with the western side of the belt lying at an altitude of about 900 m above sea level and recording mean temperatures of 24LJ, while its eastern side has a lower altitude of about 450m above sea level and higher mean temperatures of 30LJ.

### 2.3 The study design

The study utilized a cross-sectional design, with randomly selected respondents sampled in a survey carried out between July and August 2021. The number of individuals needed for the study was calculated using the standard approach for determining the minimal number of subjects needed to estimate a population proportion, n= Z_2_. p. (1 - p)/d_2_ (Pourhoseingholi et al., 2013). In order to obtain a conservative estimate, an antecedent CCHFV seroprevalence in all host types, represented by p in the formula, was assumed to be 50%. Z represented a z-score from the standard normal distribution that corresponds to 95% confidence (1.96), and d stood for the precision of the seroprevalence estimate, which was assumed to be 5%. A naïve sample size estimate of 384 subjects from each group was produced using these parameter assumptions.

Given that pastoralists and livestock clustered by households, grazing units, and ecological zones, it was expected that CCHFV seropositivity levels at the subject level would be correlated at each of these levels of aggregation. For simplicity, the study considered the household as the only level of aggregation, as the CCHFV seropositivity data at this level were expected to have the highest intra-cluster correlation coefficient. To account for this challenge, the naïve sample size for pastoralists and livestock was amplified by a design effect. The design effect (Deff) was estimated using the formula Deff=1+(b−1)ρ, where ‘b’ was the number of projected samples per herd/household and ‘ρ’ is the intra-cluster correlation coefficient (ICC), which measures the rate of homogeneity. In both pastoralists and livestock, a conservative ICC value of 0.05 was used based on the principle of using a cautious estimate when no information on the magnitude of the intra-cluster correlation coefficient at a cluster level is available (van Breukelen & Candel, 2012). A total of 242 households were therefore needed for the study assuming that up to 5 people and 20 animals would be selected from each household. This resulted in a design effect of 1.95 and an adjusted sample size of 749 for livestock. In pastoralists sample size estimation, a design effect of 1.2 was used yielding a minimum sample size of 461 pastoralists. The requirement for the minimum sample size was not enforced for wild animals because only 87 animals could be sampled in the area.

### 2.4 Sample collection, handling, and transportation

Random Geographical Coordinates (RGC) that covered the study area were generated using the QGIS software and used to select households for sampling. A household that was closest to a given RGC was selected for sampling. A pretested self-administered questionnaire was used to collect background data of each household and subjects sampled. Baseline data regarding the coordinates of the household; number of animals kept; species, age, sex and body condition scores of the animals sampled; contacts for the household head; age and gender of the human subjects (pastoralists) sampled and other potential risk factors for CCHF infection were collected using Open Data Kit (ODK) database systems.

Five animals per livestock species that were kept in selected herds including cattle, camels, sheep and goats were identified by ear-tagging for sampling using a systematic random sampling procedure. The criteria that were used to select livestock herds and animals for inclusion in the study included no recent history of any vaccination, animals without any discernible clinical syndrome and provision of informed consents from the livestock owners. Veterinarians from the Department of Veterinary Services, Isiolo County were engaged for blood sampling. They collected 10 ml of venous blood from the jugular veins from each animal.

Consenting participants, except children under the age of two years, were enrolled in the study and sampled after providing written consents. Parents or guardians provided written consent for children that were under the age of 18 years. Pastoralists sampling was done by qualified and registered nurses from the Ministry of Health. They used non-heparinized vacutainer tubes to collect up to 5 ml of blood from the median cubital vein of the left hand from each subject.

The samples obtained from both pastoralists and livestock were promptly preserved and transported to a nearby laboratory in specialized cool boxes that kept temperatures between 4 - 8°C. Upon arrival at the local laboratory the samples underwent centrifugation at 2500 ×g for 15 minutes to extract the serum. The extracted serum was then put into cryovial tubes with distinctive barcode labels. Subsequently, they were stored and transported at −20°C using a motorized freezer. The samples were also transported in this freezer to Nairobi’s International Livestock Research Institute (ILRI). The samples were brought to ILRI and put in a freezer where they stayed until they were ready for analysis at a constant temperature of −20°C.

The wild animals sampling surveys targeted various wild animals species such as giraffe, buffaloes, zebra, waterbuck, oryx, impala and warthog. Animal trapping, restraining and sampling was done by officers from the veterinary department at the Kenya Wildlife Service (KWS). All the animals were captured by chemical immobilization, except warthog that were physically captured.

Chemical immobilant comprised a combination of an opioid Etorphine 9.8 mg/mL and a tranquilizer Azaperone 100 mg/ml. This was used to dart and immobilize the target animals. Blood samples were collected from tranquilized animals using a 10ml plain vacutainer tube from the jugular vein. After sampling, the opioid antagonists – Naltrexone 50 mg/ml or Diprenophine 12 mg/ml – were used to reverse the general anesthesia. Animals were then allowed to escape as soon as they were able to wake up. Warthogs were gently nudged into linear nets for capture using the field vehicles, then physically restrained before the collection of up to 10 ml of blood from the jugular vein using plain vacutainer tubes.

The procedure for sample storage, transportation, centrifugation, labeling and processing for all the wild animals samples was similar to that used for the livestock and pastoralists samples described above.

### 2.5 Serological assay

CCHF double-antigen and multispecies ELISA Kit (ID Screen; IDvet, Grabels, France) was used to screen the serum samples collected in the study. As per the manufacturer’s instructions, the kit is 99.8% sensitive and 100% specific.

Fifty microliters of the ELISA kit dilution buffer was added to 96-well microplates well pre-coated with CCHFV recombinant N protein before 30 µl of each test sample or control were added to the specified test and control wells. The plate was then wrapped and incubated at 25°C for 45 minutes. Each test well received 50 µl of the reconstituted nucleoprotein-Horseradish Peroxidase conjugate following a washing procedure, and each well then underwent a 30-minute incubation at 25°C. Following this, 100 µl of the substrate was added after a second washing process, and the mixture was incubated at 25°C for 15 minutes. To halt the reaction, 100 µl of stop solution was added to ach well.

A spectrophotometer set to 450 nm was used to determine the optical density value (OD) for each test sample or the control. The OD obtained for the tested sample was then divided by the test’s positive control and multiplied by 100 to determine the sample-to-positive control ratio percent (S/P%). The S/P% was utilized to interpret the results; serum with a S/P% of less than or equal to 30% was regarded negative, while serum with a S/P% of more than 30% was considered to be positive (Grech-Angelini et al., 2020). Data generated from the serological assays were entered into a database that was designed using Microsoft Excel version 2018 (IBM, California).

### 2.6 Environment data

The environmental variables used in the study are outlined in S1 Table. Raster (tif) files for each of these data sets were obtained from on-line databases and clipped using the study area’s shapefile. See Supplementary S1 Table for additional details on the environmental data.

### 2.7 Statistical analysis

#### 2.7.1. Data preparation

Three types of data were available for statistical analysis for each host type (pastoralists, livestock and wild animals) sampled; these were the data collected from the field that captured independent variables that could be used for risk factor analysis, the laboratory data that characterized each subject based on their CCHFV serological status, and environment data which were also used for risk factor analysis. All the analyses were conducted in R version 4.2.2.

The first step in the analysis was to merge all the data sets by host type. The spatial coordinates collected during sampling were used to extract the environment data layers. All the pastoralists and animals sampled in the same sampling points therefore had similar values for each environment later. Vapor pressure deficit (haP) was generated as an additional environment variable using temperature and humidity values following the algorithms described by Hoch et al. (2018). Fig 2 illustrates temperature, humidity and vapor pressure deficit estimates for the study area obtained from ECMWF.

**Fig 2:**
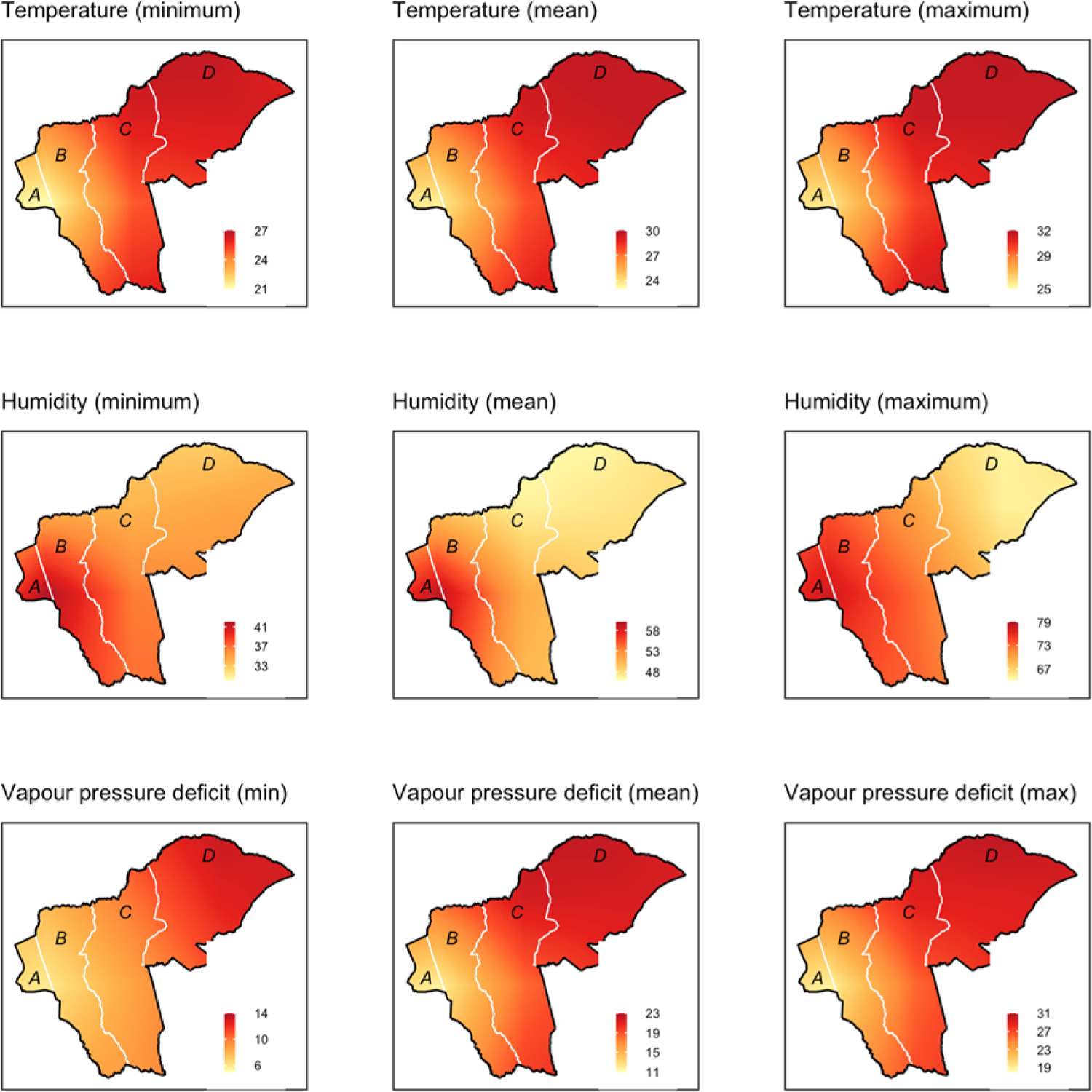
Maps showing estimates of temperature, humidity and vapor pressure deficit in the study area based on the data obtained from ECMWF. The letters A – D stand for the names of th wards which are A-Amwathi, B- Kinna, C- Garbatulla and D-Sericho

#### 2.7.2. Descriptive analyses

Descriptive analyses were conducted using the Desctools package (https://cran.r-project.org/package=DescTools). These analyses generated CCHFV seroprevalences with their 95% confidence intervals (CI), and stratified these by the independent factors determined in the field. The independent factors considered for the pastoralists data included age, gender, and the administrative ward where the subject belonged, while those that were considered for livestock included species, sex, age and ward. Given that there were few records for the wild animals, wild animals data were collapsed into two categories. The first included all the records from bovidae (buffalo, oryx, waterbuck and impala) while the others included non-bovidae non-bovidae (giraffe, zebra and warthog). Fisher’s exact tests were used to estimate unadjusted associations between the independent factors and CCHFV seroprevalence.

#### 2.7.3. Statistical modelling

Univariable and multivariable modelling was conducted to identify risk factors that influenced CCHFV seroprevalence. The univariable analysis were conducted using Generalized Linear Model (GLM). This was aimed to explore unadjusted association between the various independent factors considered and the CCHFV seroprevalence. For the pastoralists data, univariable analysis used gender, age group, ward, elevation (dem), Normalized Difference Vegetation Index (NDVI), aridity, mean annual rainfall (mrain), CCHFV seroprevalence in livestock, temperature, humidity and Vapor Pressure Deficit (VPD). Gender variable was coded as male or female and age group included 2-14 years, 15-29 years, 30-44 years and over 44 years. The subjects sampled came from three wards and the levels used in the analysis were Sericho, Garbatulla and Kinna. The rest of the factors were used as continuous variables. CCHFV seroprevalence in livestock at the household level was estimated and merged with the pastoralists data so as to investigate whether there was any significant association between the human and livestock exposure. We assumed that the effective human and livestock interactions and contacts occurred at this level.

The analysis of the livestock data used similar data as that of pastoralists. However, age groups in livestock were classified into calf/kid/lamb, young adult and adults. Calves/kids/lambs included animals that had not been weaned and were therefore being kept around the homesteads throughout the day. Young adults included animals that had not matured but had been allowed to go out for grazing with the mature animals. The species used in the analyses included goats, sheep, cattle and camels. For the wild animals data, modelling used age, species and the sex of an animal as independent factors. Age was classified as sub-adult and adults, while species were bovidae and non-bovidae.

Multivariable analyses were conducted using a generalized linear mixed model (GLMM) to account for potential clustering of data at the household level. All the continuous variables were assessed for their linearity assumption. Two-way interaction terms between independent variables were also evaluated.

## 3. Results

### 3.1. Serological assay

The study obtained a total of 580, 2,137 and 87 records from pastoralists, livestock and wild animals. The overall seroprevalence of CCHFV in pastoralists, livestock and wild animals was 7.2% (95% CI: 3.78–15.51%), 43.1% (95% CI: 30.7–50.3%) and 41.0% (95% CI: 35–85%), respectively. Table 1 provides descriptive analyses of these data by host group.

**Table 1.**
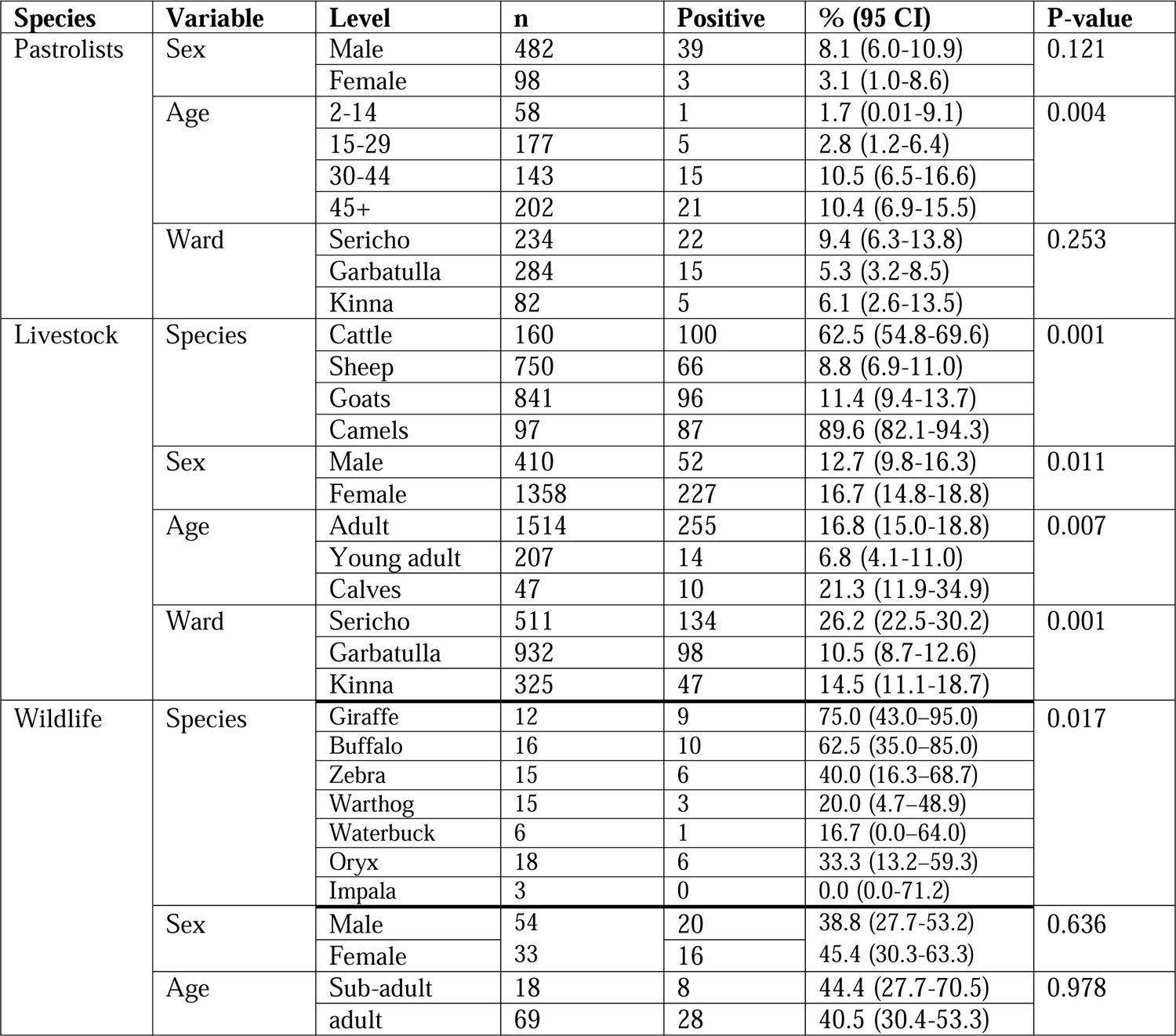
Seroprevalence of CCHFV in pastoralists, livestock and wild animals in Isiolo County.

| Species | Variable | Level | n | Positive | % (95 CI) | P-value |
| --- | --- | --- | --- | --- | --- | --- |
| Pastoralists | Sex | Male | 482 | 39 | 8.1 (6.0-10.9) | 0.121 |
|  |  | Female | 98 | 3 | 3.1 (1.0-8.6) |  |
|  | Age | 2-14 | 58 | 1 | 1.7 (0.01-9.1) | 0.004 |
|  |  | 15-29 | 177 | 5 | 2.8 (1.2-6.4) |  |
|  |  | 30-44 | 143 | 15 | 10.5 (6.5-16.6) |  |
|  |  | 45+ | 202 | 21 | 10.4 (6.9-15.5) |  |
|  | Ward | Sericho | 234 | 22 | 9.4 (6.3-13.8) | 0.253 |
|  |  | Garbatulla | 284 | 15 | 5.3 (3.2-8.5) |  |
|  |  | Kinna | 82 | 5 | 6.1 (2.6-13.5) |  |
| Livestock | Species | Cattle | 160 | 100 | 62.5 (54.8-69.6) | 0.001 |
|  |  | Sheep | 750 | 66 | 8.8 (6.9-11.0) |  |
|  |  | Goats | 841 | 96 | 11.4 (9.4-13.7) |  |
|  |  | Camels | 97 | 87 | 89.6 (82.1-94.3) |  |
|  | Sex | Male | 410 | 52 | 12.7 (9.8-16.3) | 0.011 |
|  |  | Female | 1358 | 227 | 16.7 (14.8-18.8) |  |
|  | Age | Adult | 1514 | 255 | 16.8 (15.0-18.8) | 0.007 |
|  |  | Young adult | 207 | 14 | 6.8 (4.1-11.0) |  |
|  |  | Calves | 47 | 10 | 21.3 (11.9-34.9) |  |
|  | Ward | Sericho | 511 | 134 | 26.2 (22.5-30.2) | 0.001 |
|  |  | Garbatulla | 932 | 98 | 10.5 (8.7-12.6) |  |
|  |  | Kinna | 325 | 47 | 14.5 (11.1-18.7) |  |
| Wildlife | Species | Giraffe | 12 | 9 | 75.0 (43.0–95.0) | 0.017 |
|  |  | Buffalo | 16 | 10 | 62.5 (35.0–85.0) |  |
|  |  | Zebra | 15 | 6 | 40.0 (16.3–68.7) |  |
|  |  | Warthog | 15 | 3 | 20.0 (4.7–48.9) |  |
|  |  | Waterbuck | 6 | 1 | 16.7 (0.0–64.0) |  |
|  |  | Oryx | 18 | 6 | 33.3 (13.2–59.3) |  |
|  |  | Impala | 3 | 0 | 0.0 (0.0-71.2) |  |
|  | Sex | Male | 54 | 20 | 38.8 (27.7-53.2) | 0.636 |
|  |  | Female | 33 | 16 | 45.4 (30.3-63.3) |  |
|  | Age | Sub-adult | 18 | 8 | 44.4 (27.7-70.5) | 0.978 |
|  |  | adult | 69 | 28 | 40.5 (30.4-53.3) |  |

### 3.2. Results of the univariable analyses

The univariable analyses of the pastoralists data suggest that there was a significant association between increased CCHFV seroprevalence and advanced age and higher livestock seroprevalence. In both cases, the odds of exposure increased linearly with age and livestock CCHFV seroprevalence. For livestock data, low mean rainfall and an increase in vapor pressure deficit were the two key environment variables that were associated with high CCHFV exposure. Similarly, animals that were sampled in Sericho also had higher CCHFV exposure than those sampled in the other wards. For wild animals data, none of the variables species was associated with CCHFV exposure.

### 3.3. Results from multivariable analyses

Table 2 combines the results from the multivariable analyses conducted on pastoralists, livestock and wild animals data. The results show that each model identified a unique set of variables that were identified as being significant. The model that fitted the pastoralists data identified gender, age and livestock seroprevalence as being significant while that fitted to the livestock data identified age, NDVI and vapor pressure deficit as being significant. None of the variables considered for the analysis of the wild animals data was significant.

**Table 2.** Mixed effects multivariable logistic regression analysis of the association between risk factors and CCHFV seropositivity in pastoralists, livestock and wild animals.

| Species | Variable | Level | OR (95% CI) | P-value |
| --- | --- | --- | --- | --- |
| Pastoralists | Gender | Female | 1.00 |  |
|  |  | Male | 3.38 (1.00 – 11.42) | 0.014 |
|  | Age group | 2-14 | 0.59 (0.07 – 5.22) | 0.641 |
|  |  | 14-29 | 1.00 |  |
|  |  | 30-44 | 4.72 (1.66 – 13.44) | 0.001 |
|  |  | 45+ | 4.72 (1.73 – 12.91) | 0.001 |
|  | Livestock seroprevalence (%) | <10 | 1.00 |  |
|  |  | 11-20 | 1.98 (0.79 – 4.94) | 0.132 |
|  |  | 21-30 | 1.51 (0.31 – 7.26) | 0.614 |
|  |  | >30 | 2.36 (1.06 – 5.23) | 0.041 |
| Livestock | Age | Adult | 1.00 |  |
|  |  | Calf/kid/lamb | 1.11 (0.44 – 2.81) | 0.757 |
|  |  | Young adult | 0.31 (0.16 – 0.60) | 0.001 |
|  | NDVI | - | 4.01 (1.15 – 13.94) | 0.028 |
|  | Vapor pressure deficit | - | 1.25 (1.13 – 1.39) | 0.001 |
| Wildlife | Species | Bovidae | 1.00 |  |
|  |  | Non-bovidae | 1.23 (0.49-3.12) | 0.664 |
|  | Sex | Female | 1.00 |  |
|  |  | Male | 0.69 (0.28-1.66) | 0.401 |
|  | Age | Adult | 1.00 |  |
|  |  | Sub-adult | 1.08 (0.34-3.32) | 0.891 |

The model fitted to the pastoralists data shows that being of a male gender was associated with significantly higher odds of CCHFV exposure compared to being of a female gender. Older people, especially those over 30 years, also had higher odds of exposure compared to younger ones. In fact, young people of 2-14 years were found to be a protective factor. Similarly, people from households that had livestock with >30% CCHFV seroprevalence had a higher risk of exposure compared to those that did not.

For the livestock data, young adult animals had significantly lower odds of CCHFV exposure compared to adults or calves/kids/lambs. Similarly, higher NDVI and vapour pressure deficit was associated with greater odds of CCHFV exposure.

### 3.4. Intracluster correlation coefficient

The intracluster correlation coefficient (ICC) was calculated to assess the degree of similarity among observations within the same cluster (household). ICC values were computed separately for the pastoralists and livestock datasets to account for the potential clustering effect in each population. The ICC values for CCHFV seropositivity in the pastoralists and livestock dataset were 0.25 (95% CI: 0.18 - 0.32) and 0.29 (95% CI: 0.12 - 0.34) respectively.

## 4. Discussion

This study investigated CCHFV seroprevalence and factors that influence the infection patterns of the virus in pastoralists, livestock and wild animals in an arid to semi-arid area in northern Kenya. The obtained seroprevalence rates showed significant variation among pastoralists, livestock and different wild animals species. Notably, livestock and wild animals exhibited a higher seroprevalence compared with the pastoralists whose seroprevalence was significantly low.

Seroepidemiological studies are increasingly being used to study the burden and risk factors of a wide range of infectious agents given its ability to detect both clinical and sub-clinical infections. This is particularly relevant for CCHFV given that most of the hosts that the virus infects do not show any clinical signs. It is also suspected that many human infections that occur especially in remote locations go undetected due to poor diagnostic facilities. The data obtained from this (and similar studies) are therefore invaluable in identifying hotspots that can be targeted for intensive surveys.

The study detected a low CCHFV seroprevalence in pastoralists. Being a male, having an advanced age (>30 years old) or belonging to a household that had livestock with a high CCHFV seroprevalence were associated with higher CCHFV seroprevalence compared to being a female, being younger or belonging to a household that had animals with lower seroprevalence. Similar findings were reported by Kinsella et al., 2004 and Papa, (2002). The higher CCHFV seroprevalence in males could be attributed to occupational exposure given that men were more likely to run outdoor occupational activities that could increase their contact with tick (Hoogstraal, 1979) or infectious animals. Males were more likely to be in charge of herding animals, trekking them to the markets or slaughtering animals. With respect to age, a high CCHFV seroprevalence was observed in the subjects that had more than 30 years. A similar finding was reported in South Africa where older farmers had higher prevalence of CCHF IgG antibodies compared to younger ones (Lwande et al., 2012). The higher CCHFV seroprevalence in older people could be associated with cumulative exposures over time. This is a common finding in many seroepidemiological investigations (Sargianou et al., 2013).

The study found a positive association between pastoralists and livestock CCHFV seroprevalence particularly if a livestock herd had high CCHFV seroprevalence. This observation indicates that a significant number of people acquired CCHFV infections either directly or indirectly from their own livestock (Atim et al., 2023). The significant association between livestock and human CCHFV infections highlights the potential impact of controlling the disease in livestock, for example through the use of acaricides, on human exposure. This finding also highlights the need to account for practices that are used to control tick-borne diseases in livestock while investigating CCHFV seroprevalence in humans.

There was an enormous variability on CCHFV seroprevalences between livestock species. Notably, camels had the highest seroprevalence that peaked at 89.6%. Similar exposure patterns have been described by Bouaicha Bouaicha et al. (2021) in southern Tunisia. The high seroprevalence in camels could suggest that this host plays an important role in the transmission of CCHFV (Bell-Sakyi et al., 2012). However, several countries with a history of repeated human infections, such as Uganda, do not raise camels in regions that have experienced repeated outbreaks. Some of the risk factors that have been identified in those studies include livestock (cattle) farming, collecting ticks and age of the subject (Atim et al., 2023).

The analyses further identified significant differences on CCHFV between age groups in livestock; mature animals and calves/kids/lambs had higher seroprevalences than the young adults. This finding is consistent with the work of (Spengler et al., 2016), and it can be supported by the fact that older animals might have had repeated exposures over time. Seroepidemiological data, as illustrated above, include cumulative exposures that an animal might have experienced throughout its life time as the outcome. Adult animals might have therefore had higher chances of being infected compared to young adults. Calves/kids/lambs however present a different exposure pattern – the high exposure levels in this age group could be associated with the presence of maternal antibodies in their system (Blanco-Penedo et al., 2021).

The CCHFV exposure patterns observed in livestock differed slightly from those of pastoralists in as far as they identified a few environmental variables that were significant. Previous studies by Dunster et al. (2002) in Kenya and Nabeth et al. (2004) in Senegal identified geographical factors as being important in defining the hotspots for the disease. During the elementary stages of the current analysis (before multivariable models were used), wards (administrative units) had a significant association with the outcome. Sericho ward exhibited the highest seroprevalence compared to Kinna and Garbatulla. On further analyses, it became apparent that Sericho also had the highest minimum vapor pressure deficit compared to the other wards (2). The vapor pressure deficit also became significant in the final model, indicating that selected environmental factors played a big role in making this area suitable for the virus transmission. *Hyalomma* ticks, the main vectors of CCHFV thrive well in areas with high vapor pressure deficit; this indicates that the same vector may be playing a role in the transmission of the virus in this region although more studies should be done to verify this observation.

The study also investigated several potential risk factors associated with the prevalence of CCHFV in wild animals. None of the factors investigated using multivariable models were significant. A previous study by (Obanda et al., 2021) did not also find significant association between sex and CCHFV prevalence in wild animals. Similarly, (Spengler et al., 2016) did not find any association between CCHFV exposure and age in wild animals. Nonetheless, it is important to recognize that age-related differences in susceptibility to CCHFV might vary across different wildlife species or populations, warranting additional research to better understand age-related dynamics. The differences in the infection patterns observed in wild animals and livestock data in the current study could be attributed to differences in the sampling techniques used. Livestock sampling was more structured and enabled the recruitment of animals from at least three age groups. The approach used for sampling wild animals did not provide a reliable way of recruiting animals of various age groups. The locations used for wild animals sampling were also clustered in the two national parks that were found in the extreme western region of the study area. Apart from the variability of CCHFV by species, no other variable was found to be significant.

It is also important to note the role of livestock-wild animals interaction in the spread of CCHFV. The grazing requirements of the Bovidae species tend to overlap with domesticated livestock especially the cattle, which facilitates cross-transmission of several infectious diseases and ectoparasites (Obanda et al., 2021). In this study, the average prevalence of CCHFV in wild animals’ bovidae species was higher compared to livestock’s average seropositivity, and because of the grazing overlap, wild animals likely serve as a source of infection to cattle. The role of wild animals, especially the bovidae species, on the epidemiology of CCHFV goes beyond its reservoir role and includes a potential source of direct human transmission. The illegal hunting, slaughter, and consumption of buffalo in Kenya (Kimwele, 2012) may cause CCHFV transmission to the hunters as they handle infected blood (Nabeth et al., 2004).

The analysis also identified normalized difference vegetation index (NDVI) as being a significant predictor on CCHFV exposure in livestock. The NDVI, which indicates vegetation health and density, has been shown to be significantly associated with the prevalence of CCHFV in both pastoralists and livestock. The study reveals that areas with a higher NDVI (mean = 0.59, SD = 0.18) have a significantly higher number of positive CCHFV samples in both pastoralists and livestock populations compared to areas with lower NDVI values (mean = 0.56, SD = 0.18) (T-test, P < 0.05, t-value = 42.02). This suggests that regions with healthier and denser vegetation might provide a more suitable environment for ticks, contributing to an increased risk of CCHFV transmission to both humans and livestock. This finding corroborates the work of (Dunster et al., 2002), which identified specific geographic areas as potential risk zones for CCHFV outbreaks.

The results of the environmental factors analysis highlight the significance of certain ecological parameters in influencing the prevalence and transmission of Crimean-Congo Hemorrhagic Fever Virus (CCHFV) to both pastoralists and livestock. Understanding these environmental factors is vital for developing targeted and effective strategies to mitigate the risk of CCHFV transmission and enhance disease surveillance and prevention in both human and livestock populations.

The intracluster correlation coefficient is critical in our study as it provides a quantifiable measure of the similarity among observations within the same cluster. The ICC values we got for each group (pastoralists and livestock) provide insight into the degree to which CCHFV seropositivity is clustered within specific households. This information is crucial because a high ICC indicates that individuals within the same household are more similar in terms of CCHFV seropositivity, suggesting a substantial influence of local factors or shared characteristics. Understanding this clustering effect is vital for targeted interventions, as interventions tailored to specific regions may yield more pronounced effects. Second, accurate estimate of the model parameters and reliable inference depend on the analysis taking clustering into account. Clustering can be overlooked, which can result in understated standard errors and inflated type I error rates. As a result, the addition of ICC to our study gives a more realistic depiction of the real variation in seropositivity, strengthening the robustness and reliability of our results.

While this study has yielded important insights, there are several limitations that warrant consideration in future research endeavors. Firstly, the reliance on serology introduces potential misclassification biases, which should be addressed in subsequent investigations. Additionally, the limited number of wild animals samples included in this study may restrict the breadth of our understanding of CCHFV dynamics in the ecosystem.

Furthermore, the absence of comprehensive data on specific tick species and viral isolation hinders a thorough examination of tick-vector competence and the prevailing CCHFV strains within the region. Integrating vector surveillance and viral genotyping into future investigations is crucial for advancing our comprehension of the virus’s transmission dynamics.

## Conclusion and recommendations

However, the study’s results have crucial implications for public health. The identification of high-risk areas for CCHFV transmission can inform targeted prevention and control strategies. Increasing public awareness about CCHFV transmission, especially among high-risk populations and healthcare workers, is essential for early detection and prompt management of cases. Furthermore, close collaboration between the human and animal health sectors is crucial for effective disease surveillance, outbreak response, and preventive measures.

In conclusion, this study showed that CCHFV is prevalent in the study area even though no clinical events of the disease have been reported previously. The virus is highly prevalent in livestock and wild animals, and these may serve as the source of infection to tick vectors. The study also identified NDVI and VPD as being important environment risk factors. Both of these are known to facilitate the development of ticks.

## Data Availability

All data is completely available without any restrictions. Kindly contact the corresponding author at for all data requests.

## Acknowledgements

We extend our heartfelt gratitude to the invaluable participants of our study whose cooperation and participation were instrumental to the success of our research. The dedication and hard work of our study team were essential in ensuring the smooth execution of this project. We are also indebted to the officials in the national ministries of Health and Agriculture, as well as the County government of Isiolo, for granting approval for this study to take place. We would also like to thank officers from the veterinary department at the Kenya Wildlife Service (KWS) for helping us with wild animals blood sampling.

## Supplementary data

**S1 Table.** A table showing the environment variables used in the study, including their units of measurement, source and the primary resolution.

| Dataset | Units | Source | Spatial resolution |
| --- | --- | --- | --- |
| NDVI | Index | Google Earth Engine | 30 m |
| LST | °C | Google Earth Engine | 30 m |
| Land use/land cover |  | Google Earth Engine | 30 m |
| Rainfall | Millimetres (mL) | https://data.chc.ucsb.edu/products/CHIRPS-2.0/africa_3-monthly/tifs/ | 5 km |
| Elevation | Meters | https://www.diva-gis.org/gdata). | 30 m |
| Temperature | K | ECMWF | 11 km |
| Humidity | % | ECMWF | 11 km |

## Author Contributions

Conceptualization: BB EM HMA MM JA AM.

Data curation: BB EM JA MM JA AM.

Formal analysis: EM BB DN.

Investigation: JA MM AM RN EM LJK.

Methodology: MM AM JA BB EM RN.

Visualization: EM BB.

Writing – original draft: EM BB JLB MM AM HMA JA.

